# Child neurodevelopmental risk and parental depression at 2 years in the French ELFE birth cohort

**DOI:** 10.64898/2026.05.15.26353304

**Authors:** Julie Chastang, Gladys Ibanez, Sohela Moussaoui, Nathanael Lapidus, Cecilia Sandanha Gomes, Hugo Figoni, Kim Bonello

## Abstract

Parental depression and early child neurodevelopment are closely interconnected, yet few population-based studies have examined both maternal and paternal depression in relation to early child neurodevelopmental risk. This study examined the association between child neurodevelopmental risk and parental depression in the French national birth cohort Étude Longitudinale Française depuis l’Enfance (ELFE). This cross-sectional analysis included 12,953 children and their parents who participated in the 2-year follow-up. Child neurodevelopmental risk was assessed at age 2 years using the Modified Checklist for Autism in Toddlers and categorized as low, intermediate, or high risk. Parental depression was assessed using the Kessler Psychological Distress Scale and defined as maternal depression, paternal depression, or depression in at least one parent. Multivariable logistic regression models were adjusted for sociodemographic, pregnancy-related, and child characteristics. Compared with low child neurodevelopmental risk, intermediate risk was associated with higher odds of maternal depression and depression in at least one parent. High child neurodevelopmental risk was associated with substantially higher odds of maternal depression and depression in at least one parent. Associations with paternal depression were weaker and were no longer statistically significant after adjustment. These findings suggest that parental depression, particularly maternal depression, is associated with early child neurodevelopmental risk from the stage of initial developmental concerns. They support an integrated, family-centred approach combining early identification of child developmental vulnerability with attention to parental mental health.

## Introduction

Parental mental health is increasingly recognised as a critical determinant of child and family wellbeing. Depression in parents, particularly during the perinatal period, has been consistently associated with poorer developmental outcomes in offspring, including emotional and behavioural problems, cognitive delays and impaired social functioning (1,2). Historically, research has focused mainly on maternal depression; however, emerging evidence highlights that paternal depression also exerts significant and long-lasting effects on child emotional and neurodevelopmental trajectories (3,4). The recognition that both parents play a role in shaping a child’s developmental environment underscores the need for integrated, family-centred approaches in mental health research and practice.

Beyond parental wellbeing, the child’s neurodevelopmental status is increasingly viewed as both an outcome and a determinant of family mental health. Neurodevelopmental disorders (NDDs) encompass a wide spectrum of conditions—including autism spectrum disorder (ASD), attention-deficit/hyperactivity disorder (ADHD), specific learning disorders, communication and coordination disorders, and global developmental delay—that can profoundly affect the child’s functioning and the family’s daily life (5,6). Children exhibiting early signs of developmental difficulties often require greater caregiving involvement and medical attention, which can increase parental stress and contribute to emotional distress or depressive symptoms (7). Conversely, parental depression may itself impact the child’s neurodevelopmental trajectory through genetic, prenatal, biological and psychosocial mechanisms, including altered caregiving behaviours and exposure to chronic stress (8,9).

These bidirectional and transactional dynamics between child neurodevelopment and parental mental health are increasingly documented, yet key gaps remain. Most existing studies have focused on specific diagnostic categories, especially autism spectrum disorder, while the broader continuum of neurodevelopmental vulnerabilities is less explored. Moreover, few population-based studies have simultaneously examined both maternal and paternal depression in relation to early neurodevelopmental risk, nor investigated potential moderators such as child sex, perinatal factors or socio-economic status. Evidence from large, representative European cohorts is still limited, constraining the generalisability of findings to diverse sociocultural contexts.

The present study used data from the French national birth cohort Étude Longitudinale Française depuis l’Enfance (ELFE) to examine the association between child neurodevelopmental risk, assessed using the M-CHAT, and parental depression, including maternal depression, paternal depression, and depression in at least one parent. It also examined whether this association varied according to child sex or household income.

## Methods

### Study design and population

The present study was based on data from the French national birth cohort Étude Longitudinale Française depuis l’Enfance (ELFE), which recruited newborns in metropolitan France in 2011 (10). This nationally representative cohort included 18,329 children born in 2011 in 349 maternity wards selected using random sampling methods. Mothers were ≥ 18 years old, with a single or twin live birth at ≥33 weeks’ gestation, and no plan to leave France in the following three years. The cohort was designed for long-term follow-up, up to 20 years, to examine the influence of social, environmental, and familial determinants on child health and development. Data collection began with an interview with the mother at the maternity unit at birth, followed by telephone interviews with both parents when the child was aged 2 months, 1 year, and 2 years (10).

### Variables and measurements

The outcome variable was parental depression, defined using three binary outcomes: maternal depression, paternal depression, and depression in at least one parent. Depression status was assessed when the child was approximately 2 years old, using a self-administered questionnaire completed by both parents. The presence of depressive symptoms was determined according to a predefined threshold on the selected instrument, the Kessler Psychological Distress Scale, with a threshold score of 13 or higher (11). Parents whose scores exceeded this threshold were classified as having depression. It is a reliable tool for measuring psychological distress in general and clinical populations (12).

The main exposure of interest was child neurodevelopmental risk, assessed using the Modified Checklist for Autism in Toddlers (M-CHAT), a validated 23-item screening instrument (13). The M-CHAT was administered at around 2 years of age and categorized into three levels, low, intermediate, and high risk, based on established scoring thresholds, respectively a score below 3, between 3 and 6, and 7 or higher. The low-risk category was used as the reference group.

#### Covariates

Covariates included variables potentially associated with parental depression. These comprised sociodemographic characteristics, including maternal and paternal age at the child’s birth as continuous variables, maternal and paternal educational level defined as below Bachelor’s degree or not, household income defined as belonging to the lowest quintile or not, and maternal country of birth defined as born in France or not. Pregnancy characteristics included maternal smoking during pregnancy and maternal alcohol use during pregnancy. Child characteristics at birth included firstborn status, child sex, and low birth weight, defined as a birth weight below 2,500 grams.

### Statistical analyses

Descriptive statistics were presented for each parental depression group: continuous variables as mean ± standard deviation, categorical variables as number (percentage). Differences between yes/no depression within each group were tested using Wilcoxon rank-sum test (for continuous variables) or Chi-square test (for categorical variables).

Multivariate logistic regression models were used to estimate odds ratios (OR) and 95% confidence intervals (CI) for the association between child neurodevelopmental risk (M-CHAT category) and parental depression (maternal, paternal, and depression in at least one parent). Models were adjusted for parental age, parental educational level, household income, maternal smoking during pregnancy, maternal alcohol use during pregnancy, low birth weight, child’s sex and maternal country of birth. Variables were selected a priori based on theoretical and empirical evidence of their potential role as confounders of the association between child neurodevelopmental risk and parental depression. Model fit was assessed using the likelihood ratio test, and parsimony was maintained to ensure at least 10 events per variable. All analyses were performed using R software (version 4.5.1) and statistical significance was set at p < 0.05.

### Ethical considerations

Ethical approval for data collection in maternity units and for each wave of data collection during follow-up was obtained from the national advisory committee on information processing in health research (CCTIRS: Comité Consultatif sur le Traitement de l’Information en matière de Recherche dans le domaine de la Santé) and the national data protection authority (CNIL: Commission Nationale de l’Informatique et des Libertés). The ELFE study was also approved by the National Council for Statistical Information (CNIS) (10). Parents gave their informed consent at the time of inclusion and for follow-up contacts. Access to data for this specific analysis was granted under the ELFE data access policy.

## Results

### Study population

Among the 18,040 families initially enrolled in the French national birth cohort Étude Longitudinale Française depuis l’Enfance (ELFE), 13,276 participated in the 2-year survey (Figure 1). A total of 3,841 families were excluded because the 2-year questionnaire was missing. Among participating families, 323 were further excluded because data from the M-CHAT assessment were missing. The final analytical sample included 2,953 children and their parents.

**Figure 1:**
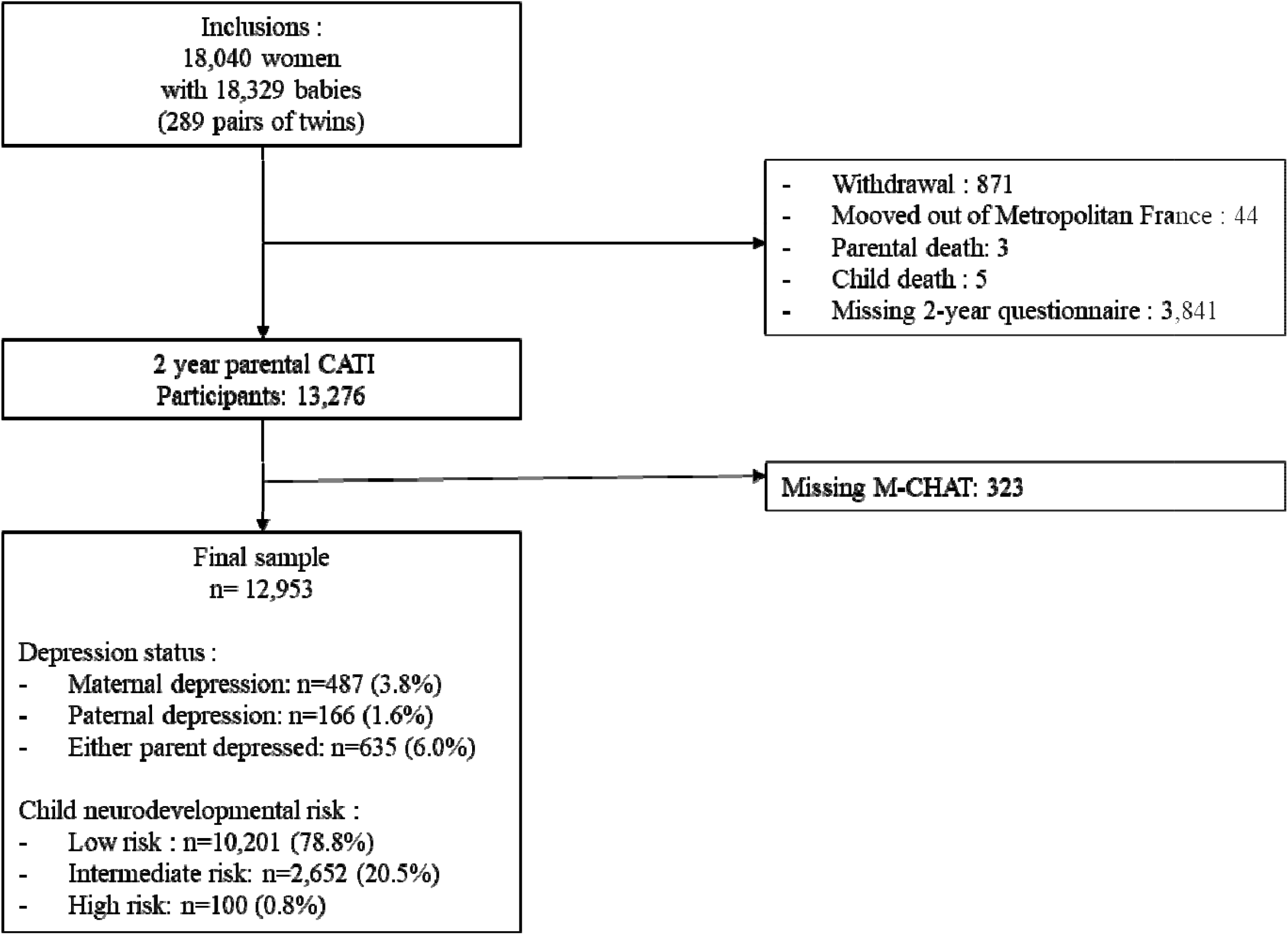
Flow chart. ELFE cohort, France, 2011-2013.

### Characteristics of the study population

Sociodemographic, pregnancy and child characteristics according to parental depression status are presented in Table 1. Overall, 3.8% of mothers and 1.6% of fathers reported depressive symptoms meeting the threshold for clinical depression, while 6.0% of families had at least one depressed parent. Mothers with depression were, on average, more likely to have a lower educational level, lower household income, and to report smoking during pregnancy. Similar but less pronounced patterns were observed for paternal depression.

**Table 1:**
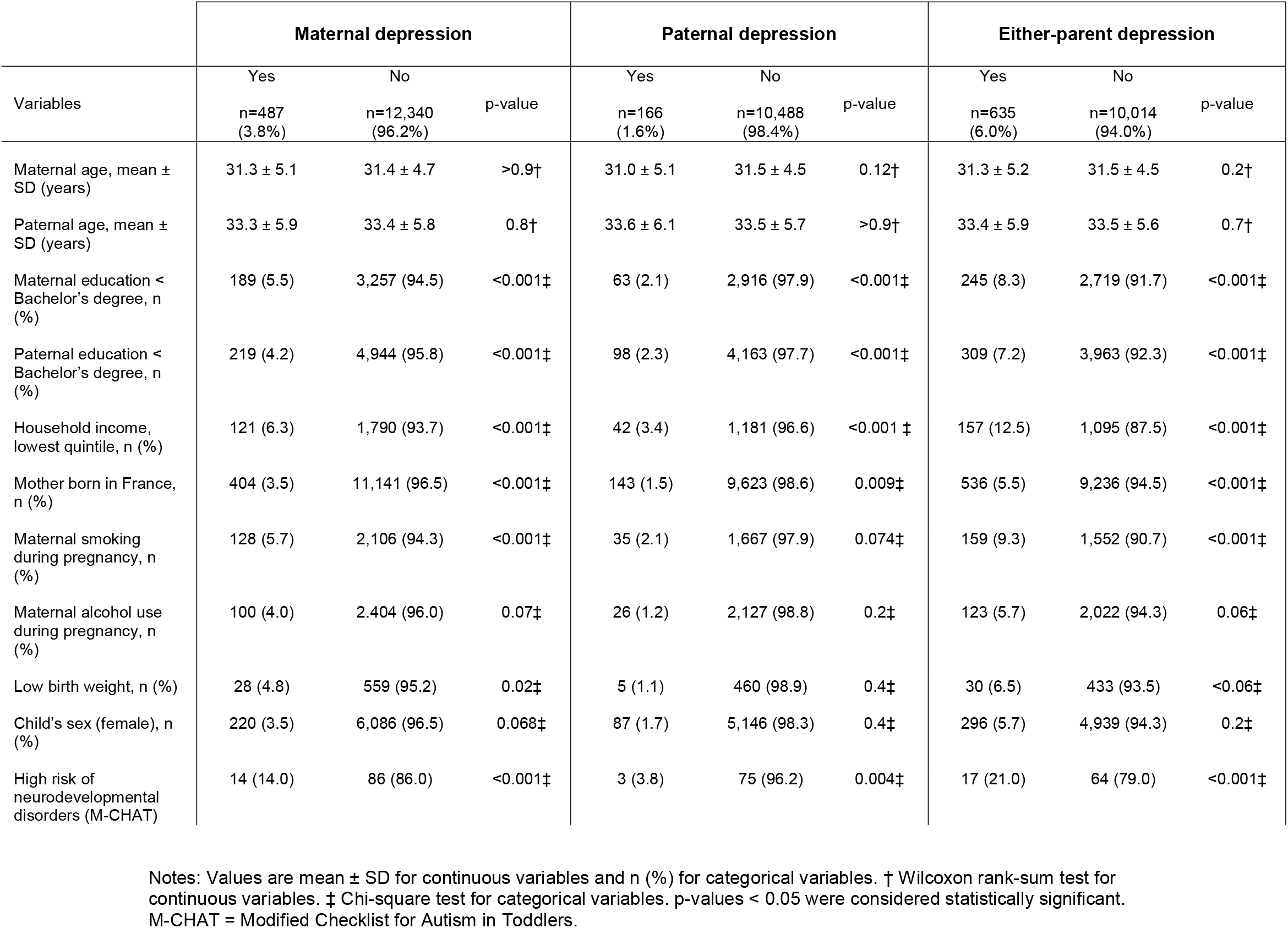
Socio-demographic, parental, pregnancy and child characteristics according to maternal, paternal, and either-parent depression status (ELFE cohort, France, 2011-2013)

|  | Maternal depression |  |  | Paternal depression |  |  | Either-parent depression |  |  |
| --- | --- | --- | --- | --- | --- | --- | --- | --- | --- |
| Variables | Yes<br>n=487<br>(3.8%) | No<br>n=12,340<br>(96.2%) | p-value | Yes<br>n=166<br>(1.6%) | No<br>n=10,488<br>(98.4%) | p-value | Yes<br>n=635<br>(6.0%) | No<br>n=10,014<br>(94.0%) | p-value |
| Maternal age, mean ± SD (years) | 31.3 ± 5.1 | 31.4 ± 4.7 | >0.9† | 31.0 ± 5.1 | 31.5 ± 4.5 | 0.12† | 31.3 ± 5.2 | 31.5 ± 4.5 | 0.2† |
| Paternal age, mean ± SD (years) | 33.3 ± 5.9 | 33.4 ± 5.8 | 0.8† | 33.6 ± 6.1 | 33.5 ± 5.7 | >0.9† | 33.4 ± 5.9 | 33.5 ± 5.6 | 0.7† |
| Maternal education < Bachelor's degree, n (%) | 189 (5.5) | 3,257 (94.5) | <0.001‡ | 63 (2.1) | 2,916 (97.9) | <0.001‡ | 245 (8.3) | 2,719 (91.7) | <0.001‡ |
| Paternal education < Bachelor's degree, n (%) | 219 (4.2) | 4,944 (95.8) | <0.001‡ | 98 (2.3) | 4,163 (97.7) | <0.001‡ | 309 (7.2) | 3,963 (92.3) | <0.001‡ |
| Household income, lowest quintile, n (%) | 121 (6.3) | 1,790 (93.7) | <0.001‡ | 42 (3.4) | 1,181 (96.6) | <0.001 ‡ | 157 (12.5) | 1,095 (87.5) | <0.001‡ |
| Mother born in France, n (%) | 404 (3.5) | 11,141 (96.5) | <0.001‡ | 143 (1.5) | 9,623 (98.6) | 0.009‡ | 536 (5.5) | 9,236 (94.5) | <0.001‡ |
| Maternal smoking during pregnancy, n (%) | 128 (5.7) | 2,106 (94.3) | <0.001‡ | 35 (2.1) | 1,667 (97.9) | 0.074‡ | 159 (9.3) | 1,552 (90.7) | <0.001‡ |
| Maternal alcohol use during pregnancy, n (%) | 100 (4.0) | 2,404 (96.0) | 0.07‡ | 26 (1.2) | 2,127 (98.8) | 0.2‡ | 123 (5.7) | 2,022 (94.3) | 0.06‡ |
| Low birth weight, n (%) | 28 (4.8) | 559 (95.2) | 0.02‡ | 5 (1.1) | 460 (98.9) | 0.4‡ | 30 (6.5) | 433 (93.5) | <0.06‡ |
| Child's sex (female), n (%) | 220 (3.5) | 6,086 (96.5) | 0.068‡ | 87 (1.7) | 5,146 (98.3) | 0.4‡ | 296 (5.7) | 4,939 (94.3) | 0.2‡ |
| High risk of neurodevelopmental disorders (M-CHAT) | 14 (14.0) | 86 (86.0) | <0.001‡ | 3 (3.8) | 75 (96.2) | 0.004‡ | 17 (21.0) | 64 (79.0) | <0.001‡ |

Notes: Values are mean ± SD for continuous variables and n (%) for categorical variables. † Wilcoxon rank-sum test for continuous variables. ‡ Chi-square test for categorical variables. p-values < 0.05 were considered statistically significant. M-CHAT = Modified Checklist for Autism in Toddlers.

Children of depressed parents were more likely to be classified as high risk for neurodevelopmental difficulties based on the M-CHAT screening. The prevalence of high M-CHAT risk was 0.6% among children of non-depressed parents and 2.7% among those with at least one depressed parent (p < 0.001).

### Association between child neurodevelopmental risk and parental depression

The associations between child neurodevelopmental risk and parental depression are summarised in Table 2. In unadjusted models, both intermediate and high risk of neurodevelopmental disorders were significantly associated with maternal and either-parent depression, while intermediate risk was also associated with paternal depression. Compared with children classified as low risk, those with intermediate risk had 1.57-fold higher odds of maternal depression (95% CI 1.28–1.92, p < 0.001), 1.65-fold higher odds of paternal depression (95% CI 1.16–2.31, p = 0.004), and 1.66-fold higher odds of either-parent depression (95% CI 1.38–1.99, p < 0.001). High neurodevelopmental risk was associated with markedly higher odds of maternal (OR = 4.71, 95% CI 2.54–8.10) and either-parent depression (OR = 4.82, 95% CI 2.72–8.11, p< 0.001), but the association with paternal depression did not reach statistical significance (OR = 2.88, 95% CI 0.70–7.87, p = 0.076).

**Table 2:**
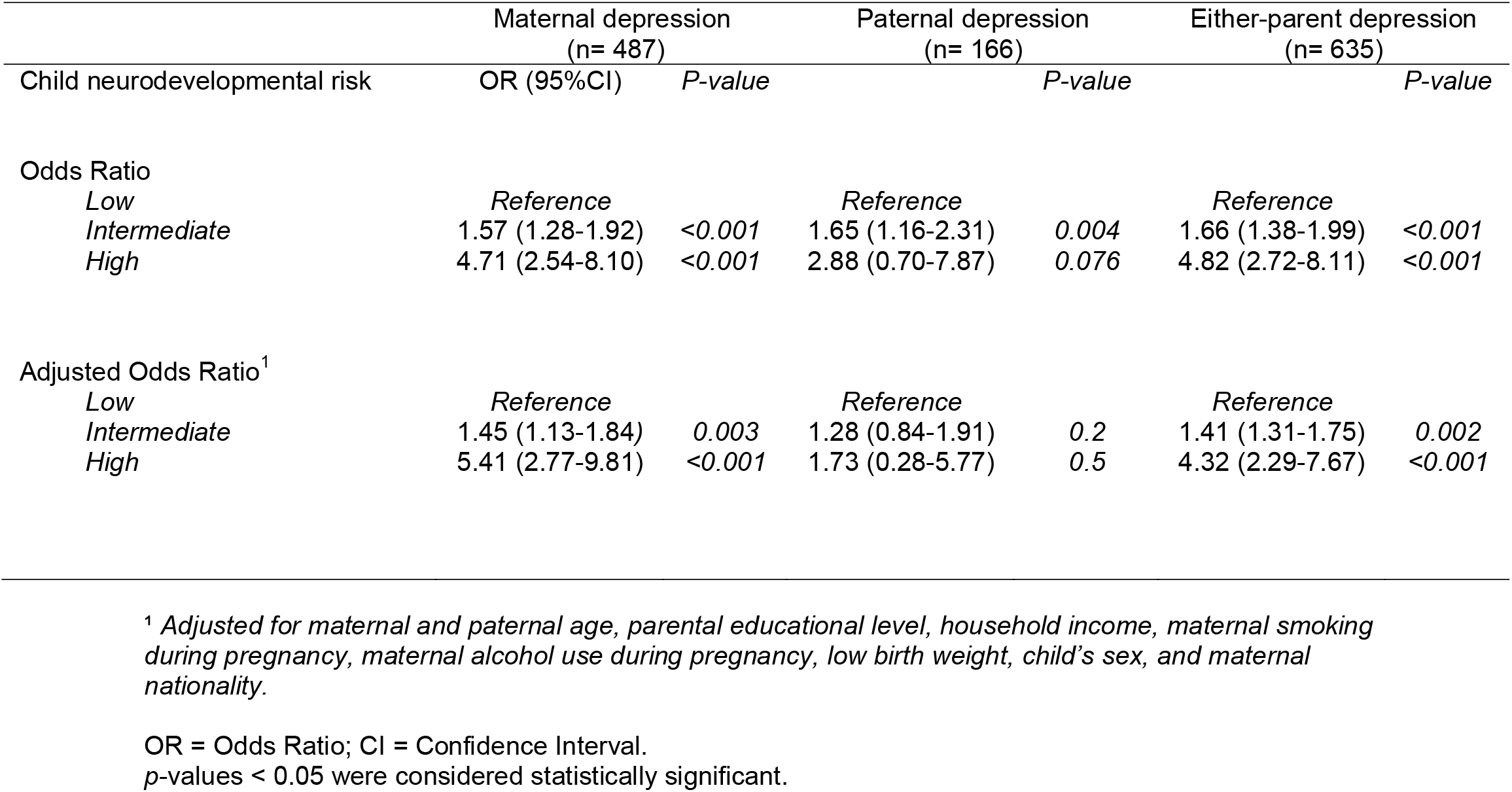
Association between parental depression and child neurodevelopmental risk (ELFE cohort, France, 2011–2013)

|  | Maternal depression<br>(n= 487) |  | Paternal depression<br>(n= 166) |  | Either-parent depression<br>(n= 635) |  |
| --- | --- | --- | --- | --- | --- | --- |
| Child neurodevelopmental risk | OR (95%CI) | P-value |  | P-value |  | P-value |
| Odds Ratio |  |  |  |  |  |  |
| Low | Reference |  | Reference |  | Reference |  |
| Intermediate | 1.57 (1.28-1.92) | <0.001 | 1.65 (1.16-2.31) | 0.004 | 1.66 (1.38-1.99) | <0.001 |
| High | 4.71 (2.54-8.10) | <0.001 | 2.88 (0.70-7.87) | 0.076 | 4.82 (2.72-8.11) | <0.001 |
| Adjusted Odds Ratio<sup>1</sup> |  |  |  |  |  |  |
| Low | Reference |  | Reference |  | Reference |  |
| Intermediate | 1.45 (1.13-1.84) | 0.003 | 1.28 (0.84-1.91) | 0.2 | 1.41 (1.31-1.75) | 0.002 |
| High | 5.41 (2.77-9.81) | <0.001 | 1.73 (0.28-5.77) | 0.5 | 4.32 (2.29-7.67) | <0.001 |

<sup>1</sup> Adjusted for maternal and paternal age, parental educational level, household income, maternal smoking during pregnancy, maternal alcohol use during pregnancy, low birth weight, child's sex, and maternal nationality.
OR = Odds Ratio; CI = Confidence Interval. p-values < 0.05 were considered statistically significant.

After adjustment for maternal and paternal age, parental education, household income, maternal smoking and alcohol consumption during pregnancy, low birth weight, child’s sex, and maternal nationality, these associations remained robust. In the adjusted model, intermediate neurodevelopmental risk remained significantly associated with maternal depression (aOR = 1.45, 95% CI 1.13–1.84, p = 0.003) and either-parent depression (aOR = 1.41, 95% CI 1.31–1.75, p = 0.002). The association was strongest for children classified as high risk (maternal depression aOR = 5.41, 95% CI 2.77–9.81, P <0.001; either-parent depression aOR = 4.32, 95% CI 2.29–7.67, p <0.001). Associations with paternal depression did not reach statistical significance.

## Discussion

This study found a significant association between child neurodevelopmental risk, as assessed by the M-CHAT at 2 years of age, and parental depression. Children classified as being at intermediate or high neurodevelopmental risk had higher odds of maternal depression and of depression in at least one parent, even after adjustment for a broad range of socio-demographic and perinatal factors. By contrast, the association with paternal depression was weaker and no longer statistically significant after adjustment. Taken together, these findings support the view that early child developmental vulnerability and parental mental health are closely intertwined and support an integrated, family-centred approach to early identification and prevention.

### Strengths and limitations

This study has several strengths. First, it is based on a large national birth cohort, which enhances the robustness of the analyses and the population relevance of the findings. Second, it examines maternal depression, paternal depression, and depression in at least one parent within the same analytical framework, providing a more comprehensive view of family mental health than studies restricted to mothers alone. Third, the analyses were adjusted for a wide range of potential confounders, thereby strengthening the credibility of the observed association between child neurodevelopmental risk and parental depressive symptoms.

Several limitations should nevertheless be acknowledged. The cross-sectional nature of the analysis does not allow the direction of the observed association to be established. Early neurodevelopmental difficulties in the child may increase parental distress, but parental psychological distress may also influence child development through biological, relational, and environmental pathways. In addition, the data relied on self-administered questionnaires, which may introduce reporting bias. Finally, the M-CHAT is a screening tool rather than a diagnostic instrument; some degree of misclassification of neurodevelopmental risk therefore remains possible, although the instrument is widely used for the early identification of children with signs suggestive of neurodevelopmental disorders (13).

### Comparison with the literature

Our findings show that the odds of parental depression increase from the intermediate level of neurodevelopmental risk onward, with a steeper gradient at the high-risk level. This point is particularly important because it suggests that the impact on parental mental health is not limited to situations in which a neurodevelopmental disorder has already been formally identified. Rather, parental vulnerability may emerge as soon as developmental concerns arise, at the stage of screening and uncertainty.

These results are consistent with the literature showing that parents of children with developmental disorders are at increased risk of depression and anxiety (1,14). A systematic review reported that approximately one third of parents of children with intellectual or neurodevelopmental disorders experience clinically significant depressive symptoms, a prevalence substantially higher than that observed in the general population (14). Our findings extend this body of evidence by suggesting that parental vulnerability may already be detectable before a formal diagnosis is made, when early signs of neurodevelopmental vulnerability are first identified.

Our results also fit with the bidirectional perspective increasingly described in the literature. A study conducted during the COVID-19 pandemic highlighted a reciprocal interaction between parental mental health and the quality of life of children with neurodevelopmental disorders, an interaction that appeared to be amplified in a context of high environmental stress (6). This transactional framework may help explain why early developmental difficulties in the child can affect parental psychological well-being, while parental distress may in turn alter early interactions, caregiving availability, and the broader family climate.

This interpretation is further supported by recent work on perinatal mental health showing that parental anxiety and depressive symptoms are associated with child neurobehavioural development through early biological, relational, and environmental mechanisms (15). Considered together, these findings support a developmental and systemic perspective in which parental and child vulnerabilities evolve interdependently rather than as isolated phenomena.

### Paternal depression

In our study, the association with paternal depression was weaker and did not remain statistically significant after adjustment, although an association was observed in the unadjusted analysis for the intermediate level of neurodevelopmental risk. This result should be interpreted cautiously. It may reflect lower statistical power due to the smaller number of fathers with depressive symptoms, but it may also suggest that paternal depression is shaped by more indirect, contextual, or relational mechanisms than maternal depression in the early postnatal period.

This interpretation is consistent with previous studies showing that early paternal depression is associated with increased emotional, behavioural, and attentional difficulties in children. A longitudinal birth cohort study found that paternal depression in early childhood was associated several years later with more behavioural problems, hyperactivity symptoms, and emotional difficulties in offspring, independently of maternal depression and socio-demographic factors. The authors also reported a cumulative effect when depression persisted over time or when both parents were affected (16).

Similarly, a recent systematic review including 26 studies reported that paternal depression occurring before, during, or after birth was associated with a higher risk of emotional, behavioural, and neurodevelopmental problems in children, including attention difficulties and autism-related outcomes (17). The mechanisms proposed are both direct, through father-child interactions and paternal engagement, and indirect, through the family environment or maternal mental health. In this context, the absence of a statistically significant adjusted association in our study should not be interpreted as evidence of no effect, but rather as suggesting a relationship that may be more mediated, more diffuse, or more difficult to detect within the present analytical framework.

These results nevertheless underscore the importance of systematically including fathers in screening and intervention strategies, alongside mothers, within a comprehensive family-centred approach.

### Screening and public health implications

These findings have several clinical and public health implications. First, they support the joint identification of parental mental health difficulties and child developmental vulnerabilities, rather than separate and compartmentalised approaches. Second, they highlight the need for early family-centred interventions. Third, they reinforce the central role of primary care in identifying at-risk situations, providing first-line support, and coordinating referral to appropriate services. In a context marked by rising mental health needs and persistent barriers to care, such integrated approaches may help prevent unfavourable long-term trajectories in both parents and children.

This perspective is reinforced by recent evidence suggesting that early identification of neurodevelopmental disorders may also benefit parental mental health. A real-world study involving more than 50,000 parents found that an early screening programme delivered through a mobile application not only enabled reliable identification of several neurodevelopmental disorders, but was also associated with an approximately 31% reduction in postnatal depression (18). Although these findings should be interpreted in light of the specific context of that intervention, they suggest that reducing parental uncertainty and facilitating faster access to care may be important levers for improving parental well-being.

Complementary evidence indicates that interventions targeting the socio-emotional well-being of parents of children with neurodevelopmental disorders are promising. A recent systematic review including 16 interventional studies highlighted the value of programmes combining psychoeducation, cognitive behavioural approaches, mindfulness, and acceptance-based strategies, with convergent effects on reducing parental stress, improving emotional well-being, and strengthening social support (19). These findings support the development of early, integrated, family-centred interventions delivered in accessible and adaptable formats, including online, hybrid, or community-based models. From this perspective, articulating parental support, child developmental care, and coordination across health and social care services appears essential to optimise long-term benefits.

## Conclusion

In conclusion, this study shows that neurodevelopmental risk identified at 2 years of age using the M-CHAT is associated with parental depression, particularly maternal depression and depression affecting at least one parent. The presence of an association from the intermediate risk level onward suggests that parental vulnerability may emerge very early, before the establishment of a formal diagnosis, at the stage of initial developmental concerns.

These findings strengthen the rationale for an integrated, family-centred approach combining early identification of child developmental difficulties, screening for parental psychological distress, and coordinated care pathways. Longitudinal studies are needed to clarify the direction of the observed associations and to improve understanding of the underlying mechanisms. In this perspective, the evaluation of early family-centred interventions should be considered a priority for both research and public health.

## Data Availability

All data produced in the present study are available upon reasonable request to the authors

